# Primary causes and direct medical cost of heart failure among adults admitted with acute decompensated heart failure in a public tertiary hospital, Kenya

**DOI:** 10.1101/2024.02.13.24302769

**Authors:** Victor M. Wauye, Chrispine O. Oduor, Felix A. Barasa, G. Titus Ngeno

## Abstract

Heart failure (HF) is a major contributor of cardiovascular morbidity and mortality globally. Despite its adverse impact on health outcomes in low- and middle-income countries such as Kenya, data on the direct medical cost of HF hospitalization is limited.

This was a prospective study conducted at Moi Teaching and Referral Hospital. Patients with HF were identified by sequential medical chart abstraction. Primary causes were extracted from echocardiogram reports and adjudicated by a cardiologist. Direct medical cost of hospitalization was derived using activity-based costing, micro-costing method, and payers’ system perspective. Drivers of overall cost were explored using linear regression models.

142 participants were consecutively recruited from September to November 2022. 51.4% were females, and the overall mean age was 54 (SD 20). The leading primary cause was cor pulmonale (CP), 28.9%; then dilated cardiomyopathy (DCM), 26.1%; rheumatic heart disease (RHD), 19.7%; hypertensive heart disease (HHD), 16.9%; ischaemic heart disease (IHD), 6.3%; and pericardial disease (PD), 2.1%. Overall direct cost of HF hospitalization was Kshs. 11,470.94 (SD 8,289.57) per patient per day, with the mean length of hospital stay of 10.1 (SD 7.1). RHD incurred the highest costs, Kshs. 15,299.08 (SD 13,196.89) per patient per day, then IHD, Kshs. 12,966.47 (SD 6656.49), and DCM, Kshs.12,268.08 (SD 7,816.12). Cost of medications was the leading driver, β = 0.56 (0.55 – 0.56), followed by inpatient fees, β = 0.27 (0.27 – 0.28) and laboratory investigations, β = 0.19 (0.18 – 0.19).

Cor pulmonale, CM, RHD and HHD were the major causes of HF. The overall direct medical cost of hospitalization was extremely expensive compared with the average monthly household income per capita in Kenya. Widespread insurance cover is therefore recommended to cushion families against such catastrophic health expenditures beside public health measures aimed at addressing primary causes of HF.

## 1 Introduction

Cardiovascular diseases (CVDs) are the most common cause of global morbidity and mortality, accounting for more than 17.9 million deaths annually in the recent years. More than 75% of these deaths occur in the middle and low income countries (1). Heart failure (HF), is one of the most common primary CVD diagnosis among hospitalized medical patients in Sub-Saharan Africa (SSA), Kenya included (2,3). The main causes of HF in SSA have been traditionally described as hypertensive heart disease (HHD), dilated cardiomyopathy (DCM) and rheumatic heart disease (RHD) and less commonly, other causes of HF such as ischemic and congenital cardiomyopathies (4,5).

Treatment of HF confers significant financial burden both to the healthcare system as well the affected families, being largely driven by direct medical costs of hospitalization (6,7). Direct medical costs accounted for about 60% of the total costs in a review by Cook et al. (6). There is however limited data published on the cost of treatment of HF in Africa. Ogah et al. reported a total cost of USD 2,128 per patient per year in Nigeria, using a broader societal costing approach (8).

To the best of our knowledge, there is no published study on medical cost of hospitalization of HF in Kenya despite the efforts towards the achievement of universal health coverage. According to the 2023 Kenya Demographic Health Survey, healthcare expenditure in Kenya is mainly paid for out of pocket, with only 26.5% of the Kenyans having any form of health insurance (9). Of these, only 24% are covered by the government sponsored national health insurance fund (NHIF), while the rest are covered by private insurance. At tertiary government health facilities, about 35% of the hospitalized patients have NHIF cover (10). Furthermore, it is notable that about 83% of workforce in Kenya is comprised of informal sector (11). A study assessing the uptake of NHIF among informal workers in Western Kenya showed that only 12% had health insurance coverage, with the majority experiencing catastrophic health expenditure (12), thus implying that patients with HF would be at a high risk of financial catastrophe.

In our study, we sought to further develop the understanding of the cost of HF treatment in a low-middle-income economy, by describing the primary causes of HF at Moi Teaching and Referral Hospital (MTRH), Kenya, and determining the direct medical cost of hospitalization among patients hospitalized with acute decompensated HF (ADHF).

## 2 Methodology

### 2.1 Study design and population

This was a prospective study conducted at the Moi Teaching and Referral Hospital (MTRH), Kenya, over a period of three months from 01 September to 30 November 2022. MTRH is a national referral hospital located in Western Kenya, with a catchment area of about 24 million people. It has two wings: public and private wing, with the former being where the general public seek treatment. All those who were hospitalized to the public wing medical wards and cardiac care unit (CCU) with the diagnosis of ADHF based on the Modified Framingham clinical criteria for HF and were aged 18 years and above were included in the study. Those who were less than 18 years old, readmitted during the study period and discharged to or from the MTRH private wing were excluded from the study.

### 2.2 Case definition for the primary causes of HF

Comprehensive clinical data including clinical symptoms and signs, past medical history, and results of laboratory as well as imaging investigations were gathered and collated with the two-dimensional doppler echocardiography (ECHO) and 12-lead electrocardiography (ECG) findings. Both electrocardiograms and echocardiograms were performed by well-trained ECHO/ECG technicians at MTRH ECG/ECHO centre, according to the American Society of Cardiology guidelines (13). The findings were summarized by the principal investigator, and further confirmed by the consultant cardiologist. The most likely primary cause of HF was then established using a predetermined case definition criteria (Table 1), which was guided by the European Society of Cardiology guidelines (14). A similar criteria was previously applied in the Heart of Soweto study in South Africa (2,15), and the Sub-Saharan Africa Survey of Heart Failure which recruited participants from 9 African countries, Kenya included (4). The criteria was however tweaked to fit our set up with limited diagnostic resources.

**Table 1:**
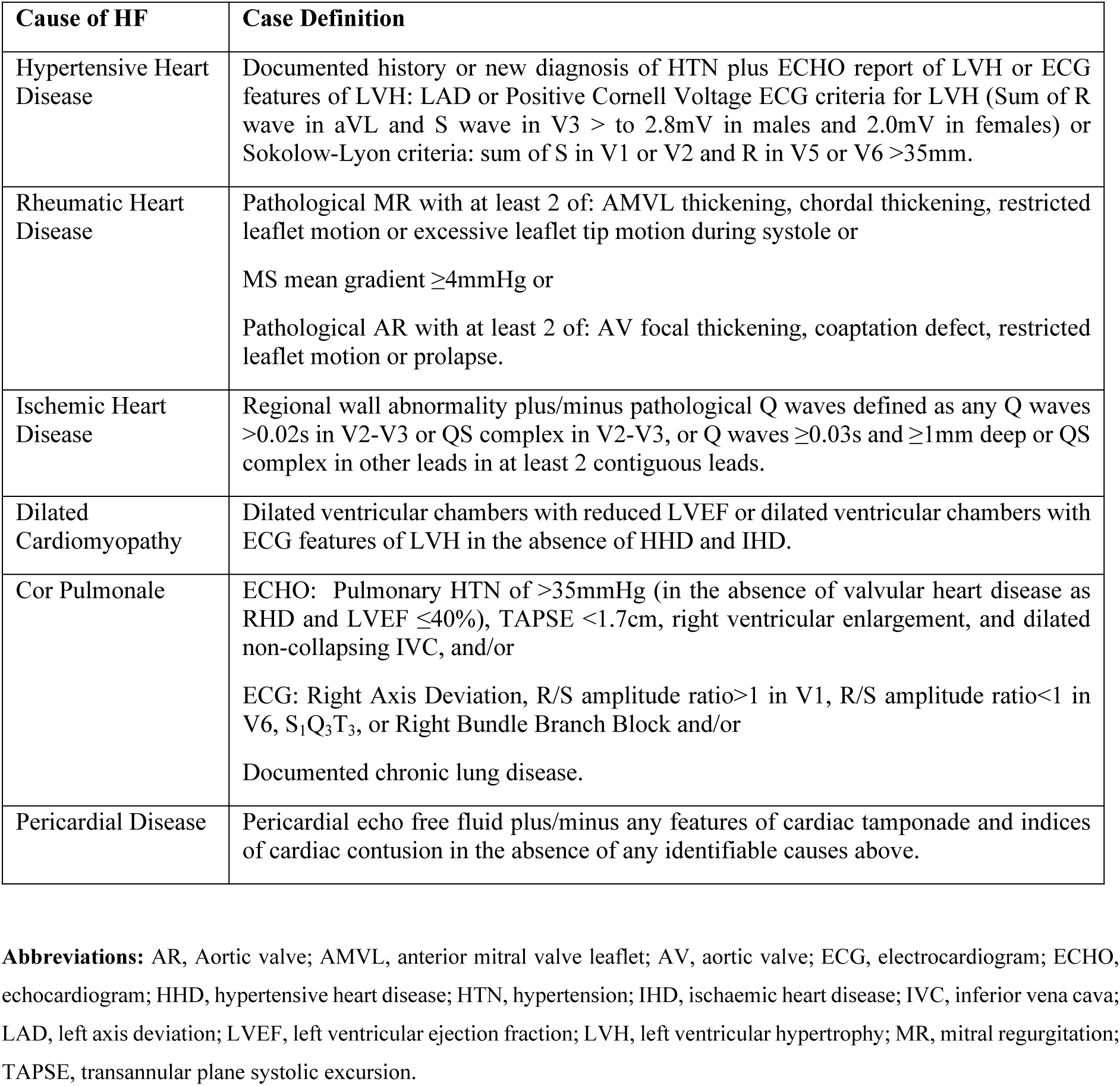
Case definition for the primary causes of heart failure.

### 2.3 Direct medical costing approaches

Costing was done using payers’ perspective(16), with a prevalence and prospective methods

(17). Detailed data regarding direct medical costs were obtained from the MTRH finance department in printed forms. The data included a breakdown of all the costs that each patient incurred during hospitalization, from the date of admission to the point of discharge (the date on which the attending physician decided that the patient be discharged, or the patient died). Micro-costing technique was used to detail the cost components, which included costs related to inpatient charges (nutrition, admission, physiotherapy, bed and nursing charges), medications, laboratory investigations, imaging, oxygen therapy, and other utilities (consumables such as syringes, gauzes, needles, nasal prongs among others) (18,19). Further, direct costs included both out of pocket (OOP) and national health insurance (NHIF) payments. Notably, being a public health facility with human resource remuneration largely covered by the government, charges related to remuneration were not included. Moreover, indirect costs such as transport and productivity losses were not included.

### 2.4 Data collection

Data were collected using interviewer administered questionnaires, and included information on socio-demographic characteristics, past medical history, New York Heart Association assessment (NYHA), vital signs such as blood pressure and heart rate, ancillary laboratory investigations such as haemoglobin, electrolytes, urea and creatinine, lipid profile, thyroid function tests, liver function tests and international normalizing ratio, chest Xray, select ECG and ECHO findings. Comorbidities were physician-diagnosed and documented in the files, largely based on the patients’ past medical history and laboratory investigations.

### 2.5 Statistical analysis

Data were analysed using RStudio statistical software version R.4.2.2. Continuous variables were reported using mean and median with respective standard deviations (SD) and interquartile range (IQR). Primary causes of HF were reported using proportions and percentages, with respective 95% confidence intervals (CI). Costs were reported using mean (SD) as cost (in Kenya shillings) per patient per day; calculated by dividing the total cost during hospitalization by the mean length of hospital stay (LOS) by the total number of participants, N, and compared between the CCU and the general ward. Linear regression model with standardized β-coefficients was used to determine how each cost component drove the overall cost. Due to inability to delineate direct medical cost of comorbidities, the mean costs were all-inclusive. However, the effect of each comorbidity on the overall cost was explored using generalized linear regression model with gamma distribution and log-link function. Regressions involved both bivariate and multivariate analyses. Comparisons of mean costs between general ward and CCU was done using Mann-Whitney and Kruskal-Wallis non-parametric tests of association. Further, sub-analysis of the overall costs was done by NYHA, grade of ejection fraction, and in-hospital outcome (discharged alive or dead). Costs were further converted to American dollars (USD) per the Central Bank of Kenya foreign exchange rates during the study period, for which 1USD = Kshs 122.6129(20).

### 2.6 Ethics statement

A formal written and signed informed consent was obtained from the participants before enrolment into the study. The informed consent described the details of the study, detailing both the benefits and potential harms. Participants’ data were decoded anonymously for confidentiality. Further, the study was approved by the Moi University Institutional Research Ethics Committee (approval number 003738 and reference number IREC/2020/187).

## 3. Results

A total of 142 participants were included in the study, after 178 cases were screened for eligibility (Fig 1). The mean age was 54 (SD 17) years, with the median age of 60 (IQR 37.3, 70). 51.4% were females. Further, 73.9% had unskilled labour, and only 52.8% had NHIF cover. The mean length of hospital stay was 10.1 (SD 7.1) days. Table 2 shows further socio-demographic, laboratory, comorbidities, medications and ECG/ECHO findings.

**Fig 1.**
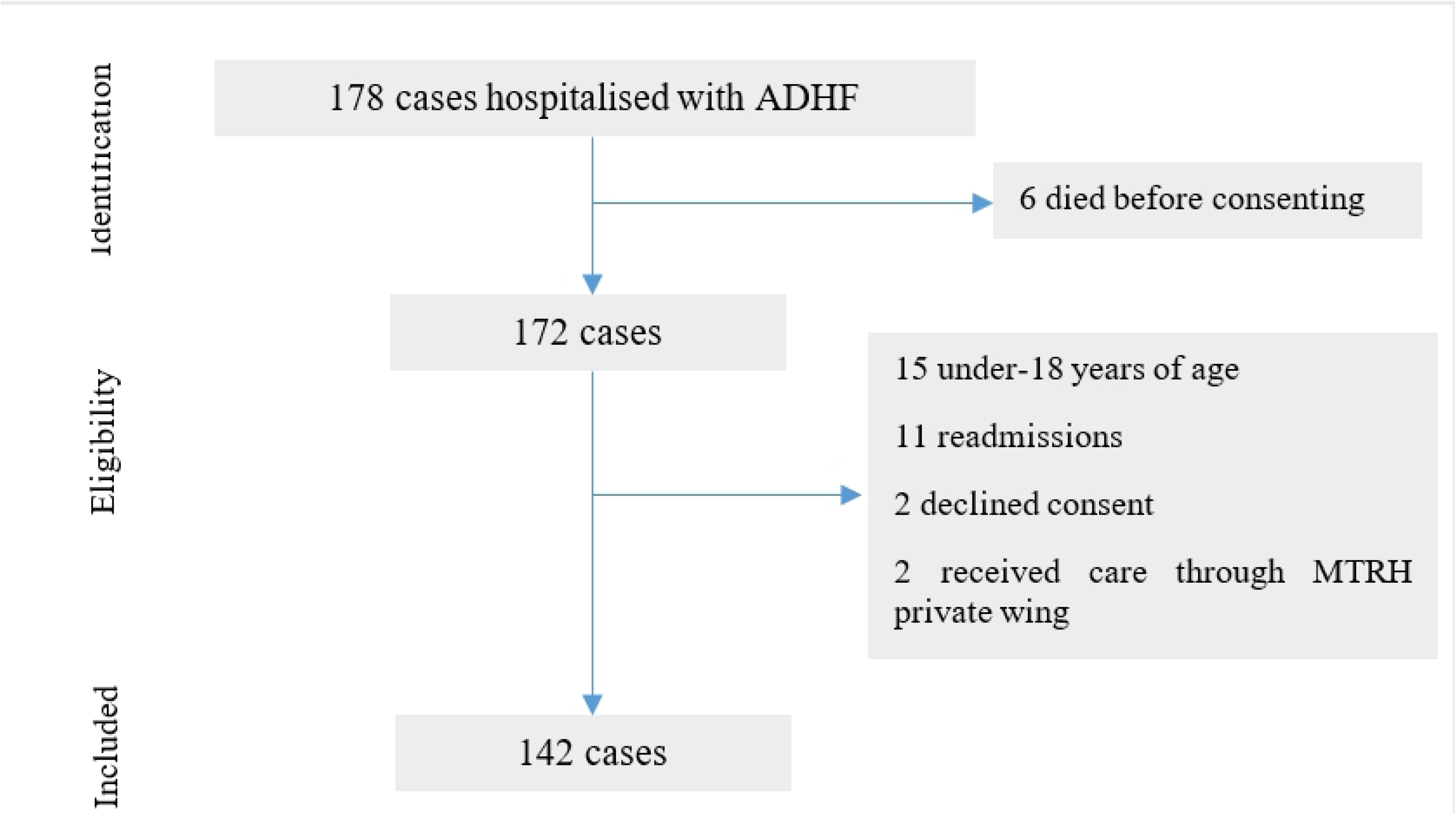
Recruitment schema of the participants

**Table 2:**
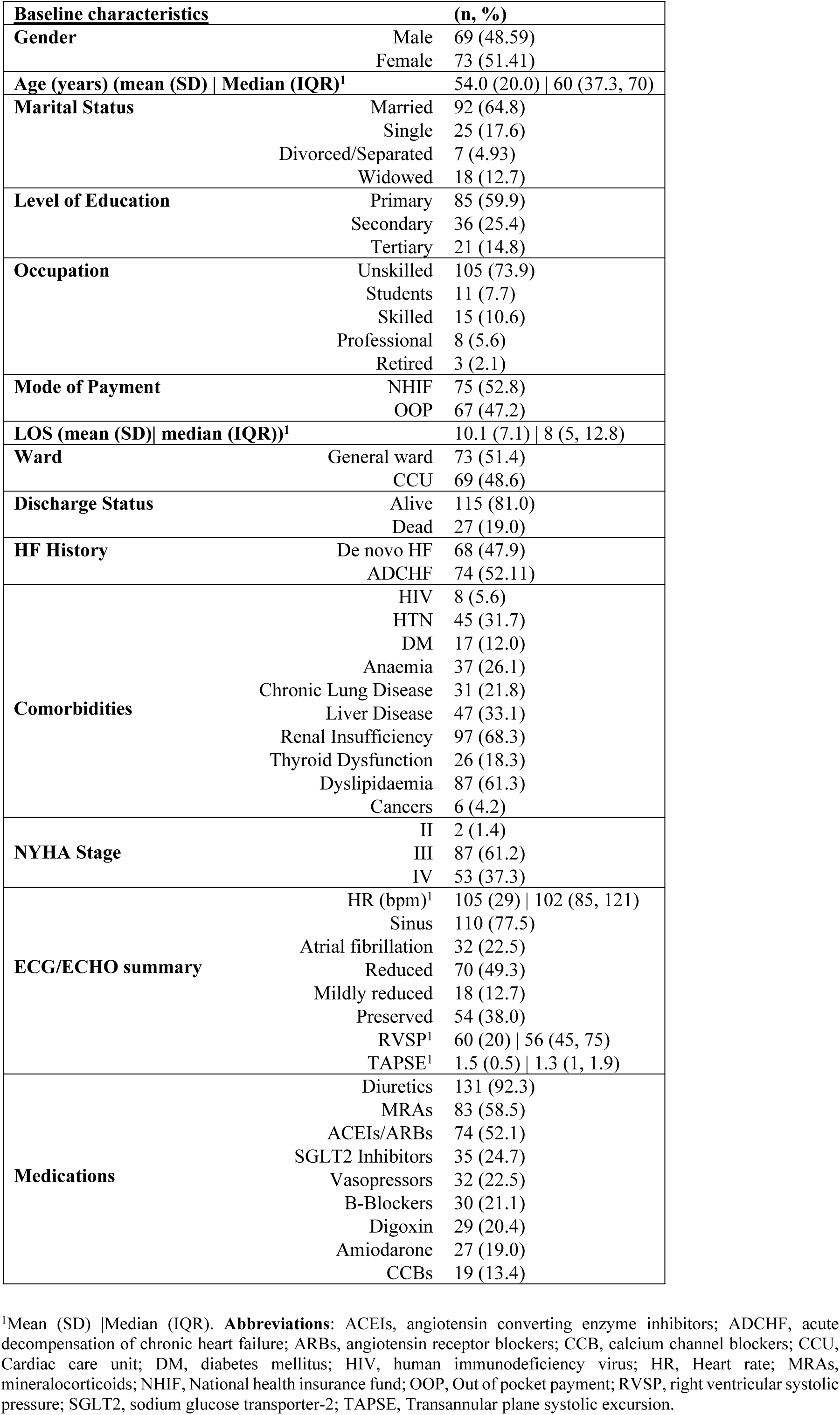
Baseline characteristics of the participants.

### 3.1 Primary causes of HF

The most common primary cause of HF was cor pulmonale (CP), accounting for 28.9% (95% CI 21.1% – 37.9%) of the cases, followed by dilated cardiomyopathy (CM), 26.1% (95% CI 18.3% - 35.0%), rheumatic heart disease (RHD), 19.7% (95% CI 12% - 28.7%) and hypertensive heart disease (HHD), 16.9% (9.2% - 25.9%) (Table 3).

**Table 3:**
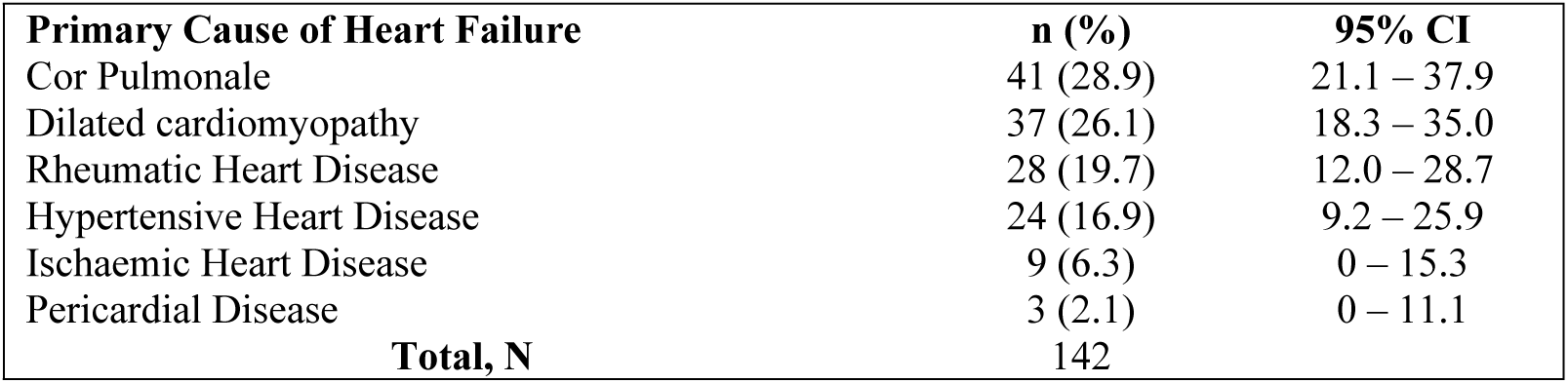
Primary causes of heart failure.

| Primary Cause of Heart Failure | n (%) | 95% CI |
| --- | --- | --- |
| Cor Pulmonale | 41 (28.9) | 21.1 – 37.9 |
| Dilated cardiomyopathy | 37 (26.1) | 18.3 – 35.0 |
| Rheumatic Heart Disease | 28 (19.7) | 12.0 – 28.7 |
| Hypertensive Heart Disease | 24 (16.9) | 9.2 – 25.9 |
| Ischaemic Heart Disease | 9 (6.3) | 0 – 15.3 |
| Pericardial Disease | 3 (2.1) | 0 – 11.1 |
| **Total, N** | **142** |  |

### 3.2 Direct medical cost of HF hospitalization

The mean direct medical cost of HF hospitalization was Kshs. 11,470.94 (SD 8,289.57) per patient per day, with the cost in the CCU being as twice as the cost in the general ward (table 4). Of the cost components, cost of medications was the leading driver of the overall cost, with standardized β-coefficient of 0.56 (95% CI 0.55 – 0.56), followed by the cost of inpatient fees and laboratory investigations respectively (Fig 2). Stratified by the primary cause of HF, RHD incurred the highest mean direct medical cost, Kshs. 15,299.08 (SD 13,196.89) per patient per day (Fig 3). Overall, direct medical cost of HF hospitalization was increasingly higher among those with NYHA stages III and IV, HFrEF and HFmrEF, comorbid renal and liver diseases, and those who died during hospitalization (S Table 1, S Table 2, S Table 3 and S Table 4, and S Fig 1).

**Fig 2.**
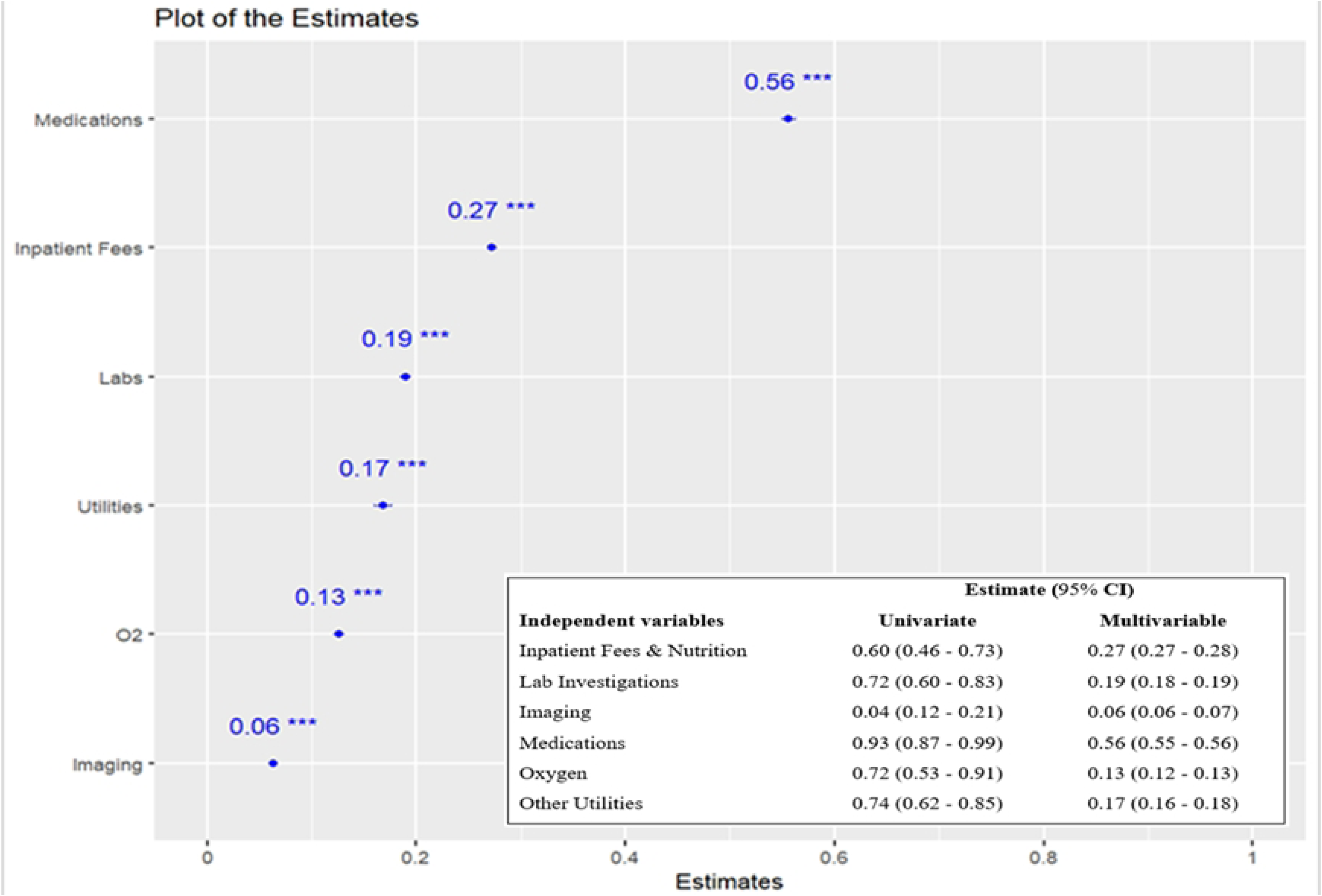
Drivers of the overall direct medical cost of HF hospitalization

**Fig 3.**
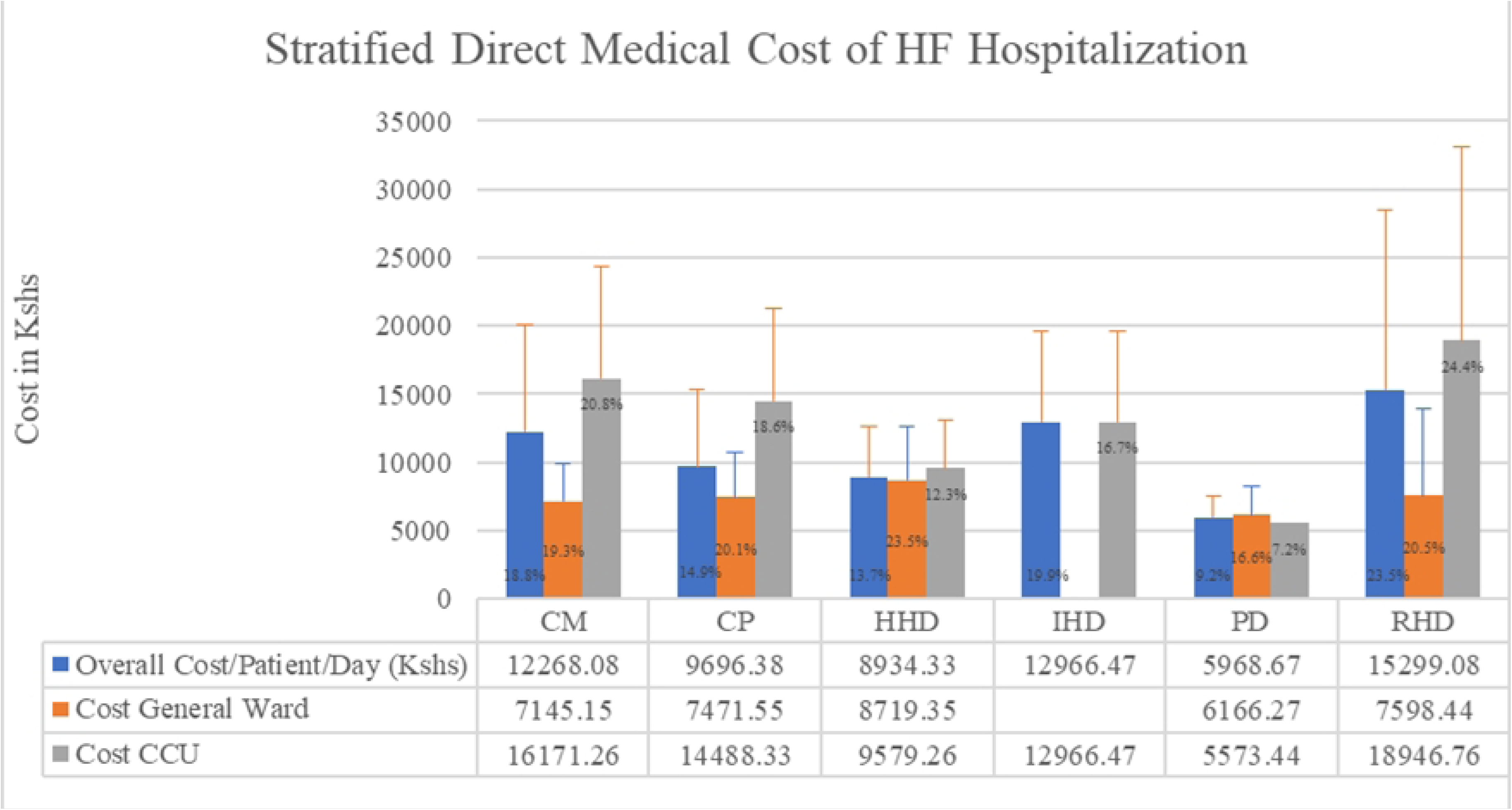
Stratified direct medical costs by the primary cause of HF

**Table 4:** Direct medical cost of heart failure hospitalization.

| Item Line | Amount (Kshs) (Mean, SD) |  |  |  |
| --- | --- | --- | --- | --- |
|  | Overall | General ward | CCU | P-value |
| Inpatient Fees | 18091.72 (20861.32) | 7919.78 (4856.96) | 28853.34 (25471.93) | <0.0001 |
| Lab Investigations | 22733.39 (14135.5) | 20953.01 (11167.76) | 24619.99 (16590.92) | 0.5124 |
| Imaging | 5139.44 (4928.71) | 6434.25 (5814.52) | 3769.57 (3296.85) | 0.0004 |
| Medications | 36694.92 (42403.40) | 18833.61 (19719.78) | 55591.67 (51105.17) | <0.0001 |
| Oxygen | 10044.44 (9743.60) | 7960.53 (6530.91) | 12373.53 (12076.73) | 0.167 |
| Other Utilities | 11427.48 (12925.69) | 7505.31 (4583.15) | 15577.02 (17035.38) | 0.0011 |
| **Cost/patient (Kshs)** | **99065.65 (76339.71)** | **65775.94 (36922.10)** | **134285.20 (90510.95)** | **<0.0001** |
| **Total cost/patient (USD)** | **807.44 (622.21)** | **536.11 (300.94)** | **1094.50 (737.71)** |  |
| **LOS (days)** | **10.1 (7.1)** | **10.0 (6.6)** | **10.3 (7.7)** |  |
| **Cost/patient/day (Kshs)** | **11470.94 (8289.57)** | **7687.57 (3783.92)** | **15473.63 (9782.79)** |  |
| **Cost/patient/day (USD)** | **93.49 (67.56)** | **62.66 (30.84)** | **126.12 (79.74)** |  |

## 4. Discussion

This study found that CP was the most common primary cause of HF among patients hospitalized with ADHF in MTRH, accounting for 28.9%. Other leading causes of HF were HHD, DCM and RHD, consistent with several previously published studies in Sub-Saharan Africa (SSA) (4,15,21–23). Our findings highlight the importance of CP, as well as the role of chronic lung diseases (CLDs), alongside other drivers of HF morbidity in SSA (24–26). Notably, 46.3% of those who had CP in this study had chronic obstructive lung disease, 24.4% had indeterminate pulmonary hypertension, 17.1% had post-tuberculosis lung fibrosis and bronchiectasis, 7.3% had lung masses and 4.9% had pulmonary embolism. Although we did not collect data on indoor air pollution, it is possible that indoor air pollution contributed to the high prevalence of chronic lung diseases hence CP. Recent studies conducted in Western Kenya reported significantly high levels of indoor air pollution due to biomass fuel use (27,28), and this was strongly associated with different cardiac abnormalities, differentially affecting women and those with low socioeconomic status (29,30). It is notable that more women than men had HF in this study; 51.4% vs 48.6%, had CP; 56.1% vs 43.9%, and 73.9% overall had unskilled labour, reflecting low socio-economic status.

This study found that the mean direct medical cost of HF hospitalization at MTRH was Kshs. 11,470.94 (SD 8,289.57) {USD 93.49 (67.56)} per patient per day. Kwok et al., in the US reported a cost of USD 11,845 (SD 22,710)(31), while Zaour et al., in Canada reported USD 10,123 per patient per hospitalization (32). Other studies from observational cohorts in SSA have estimated the of societal cost impact of HF hospitalization to be USD 2,128 per patient per year (8). Comparatively, these costs are much higher than what we found, highlighting the importance of healthcare service delivery context in cost modelling. It is possible that higher healthcare resource utilization in the developed countries, alongside different costs of health system inputs, variable health insurance coverage, and heterogeneity in the study designs resulted in the differences observed (33–36). In addition, our cost estimates represent costs in a government-subsidized health care system wherein, indirect costs as well as the human resource costs are not charged directly to patients.

The 2022 Economic Survey by the Kenya National Bureau of Statistics showed that the gross national income per capita in 2021 was Kshs. 20,122.23 (USD 164.01) per month(37). We were not able to get data on the average monthly income of the participants in this study, but it is notable that 83.5% of the participants did not complete high school, and 73.9% were unskilled labourers. Therefore, our finding of Kshs 11,470.94 (USD 93.49) indirect costs per patient per day would be extremely high against the Kenyan gross national income per capita, posing a high risk of financial catastrophe to the affected family members and public healthcare system. Although higher proportion of patients (52.8%) had national health insurance cover compared to the national average (26.5%), the higher coverage rate likely represents increased awareness of the importance of insurance cover and utilization among patients with chronic disease seeking tertiary facility care. This notwithstanding, there remains a need to intensify advocacy for and implementation of widespread national health insurance coverage among patients with HF towards the achievement of universal health coverage (UHC) and sustainable development goal (SDG) 3.8)(38,39). Further, there is need to intensify primary management and prevention strategies among patients with HF to reduce chances of hospitalization hence the high direct medical costs of hospitalization.

To the best of our knowledge, this was the first study in the region to stratify direct medical cost of HF hospitalization by aetiology. This study found that treatment of HF caused by RHD was the most expensive, costing Kshs 15,299.08 per patient per day. Sub-analysis showed that of the patients with RHD, 67.9% were admitted in the cardiac intensive care unit (CCU). RHD associated HF accounted for 27.5% of the total CCU admissions. Further, 28.6% of those admitted with RHD associated HF died, accounting for 29.6% of all those who died (S Table 5). Notably, direct medical cost of those who died was twice the cost of those who were discharged alive. Finally, RHD accounted for 37.8% of all the patients with atrial fibrillation in this study. It is possible that additional treatment modalities such as cardioversion and anticoagulation among patients with RHD and atrial fibrillation (40) increased the cost of treatment. It is further arguable that most patients with RHD were critically ill at presentation, were admitted to the CCU for intensive care, and a significant number died, driving the cost upwards. Further studies are needed to characterize costs attributable to RHD, as well as community-based prevention measures noting that RHD is a preventable disease.

The cost of medications was the leading driver of the overall cost as has been evidenced in prior literature (41,42). The overall cost also expectedly increased with advanced NYHA (S Table 1), EF (S Table 2), comorbidities particularly (S Table S, and S Fig 1), and discharge status; overall cost of those who died was as twice as that of those who were discharged alive (S Table 3). Additionally, overall treatment cost among those who were hospitalised in CCU was twice as high as treatment cost in the general wards (Table 4). This would be because of increased resource utilisation following intensive care for critically ill patients with advanced stages of HF (43–49).

This study however had the following limitations. First, participants were recruited from a public healthcare facility, hence the findings regarding cost may not be applicable to private healthcare facilities. Second, the utility of the findings on cost in designing healthcare budgets should be taken with caution because this study was conducted over a short period of time, and it may be difficult to predict how the cost may change over a longer time horizon. However, the costs reported per patient per day are intuitive and reflects daily consumption of healthcare resources during hospitalization with ADHF. Finally, payer’s perspective used for costing in this study could be narrow and may not reflect the total cost of HF treatment during hospitalization.

## 5. Conclusion

In conclusion, cor pulmonale was the most common primary cause of HF, followed by CM, RHD and HHD. The mean direct medical cost of hospitalization was Kshs. 11,470.94 per patient per day. Weighted against Kenyan monthly gross national income per capita, this was extremely expensive. Stratified by primary cause of HF, RHD incurred the highest direct medical costs. Therefore, policies and community-oriented interventions should be put in place for early recognition and treatment of CP, among other primary causes of HF in view of the ongoing epidemiological transition in SSA since these causes are completely preventable. Further studies are also recommended exploring causal relationship between wider causes of HF in SSA, particularly with the current public health challenges of global concern such as climate change and air pollution. Additionally, widespread and improved health insurance coverage should be implemented to protect the affected families and the healthcare system from catastrophic spending resulting from high direct medical cost of HF hospitalization amid the efforts towards the achievement of universal health coverage in Kenya. Finally, more extensive cost of illness studies such as societal perspective are recommended, detailing lost productivity among patients hospitalized with HF.

## Data Availability

Data used in this study is available for sharing upon reasonable request to the corresponding author.

## Acknowledgements

The authors thank all the staff and study participants for their co-operation during the study.

## Supporting information

**S Table 1. Direct medical cost by NYHA**

**S Table 2. Direct medical cost by EF**

**S Table 3. Direct medical costs by outcome status**

**S Table 4. Effect of comorbidities on the overall cost**

**S Table 5. Baseline Characteristics per primary cause of HF**

**S Fig 1. Coefficient plot of the effect of the comorbidities on the overall cost**

